# Gaps in maternal, newborn, and child health research: a scoping review of 72 years in Ethiopia

**DOI:** 10.1101/2021.01.16.21249929

**Authors:** Grace J Chan, Jenna Daniel, Misrak Getnet, Matthew Kennedy, Ronke Olowojesiku, Bezawit M Hunegnaw, Sarah Unninayar, Lisanu Taddesse, Delayehu Bekele

## Abstract

**Objectives:** Despite significant reductions in maternal and child mortality over the past few decades, a disproportionate number of global deaths occur in low and middle-income country settings, such as Ethiopia. To prioritize research questions that would generate policy recommendations for better outcomes, we conducted a scoping review that gathers the current knowledge of maternal, newborn, and child health (MNCH) and illustrates remaining gaps in Ethiopia.

**Design:** Scoping review.

**Methods:** We conducted a search strategy from 1946-2018 in PubMed/MEDLINE, EMBASE, and the WHO African Index Medicus. The study team of reviewers independently screened titles, abstracts, and full-texts; abstracted data; and reconciled differences in pairs. Descriptive analyses were conducted.

**Results:** We identified 7,829 unique articles of which 2,170 were included. Most MNCH publications in Ethiopia (70.0%) were published in the last decade, 2010-2018. Most studies included children aged one to less than 10 years old (30.5%), women of reproductive age (22.0%), and pregnant women (21.9%); fewer studies included newborns (7.0%), infants (6.6%), and postpartum women (2.9%). Research topics included demographics and social determinants of health (43.4%), nutrition (15.3%), and infectious diseases (13.0%). There were limited studies on violence (1.4%), preterm birth (0.8%), antenatal/postpartum depression (0.7%), stillbirths (0.1%), and accidents (0.1%). Most study designs were cross-sectional (53.6%). A few study designs included prospective cohort studies (5.5%) and randomized control trials (2.3%).

**Conclusions:** This is the first scoping review to describe the landscape of MNCH research in Ethiopia. Understanding the depth of existing knowledge will support the prioritization and development of future research questions. Additional studies are needed to focus on the neonatal, infant, and postpartum populations as well as preterm and stillbirth outcomes.

**STRENGTHS AND LIMITATIONS OF THIS STUDY:**

- To our knowledge, this is the first published review to have comprehensively described or summarized this body of literature in Ethiopia, allowing for a deeper understanding of the expanse of existing knowledge.
- A quality assessment of each study was not feasible due to the broad range of included topics; however, one may be conducted in the future to further refine results.
- Although conclusions may not be widely generalizable beyond Ethiopia, this review may inform the need to conduct similar reviews for all sub-Saharan countries to support effective prioritization of future research questions.

## INTRODUCTION

Globally, tremendous progress has been made in the reduction of maternal and child mortality since the turn of the century, particularly in regions facing the greatest barriers to health and development. In sub-Saharan Africa since 2000, the maternal mortality rate has declined 35% and under-five mortality rate has declined 49%.[1,2] However, despite these improvements, maternal and child mortality remains high. Of the estimated 295,000 global maternal deaths in 2017, approximately 66% occurred within sub-Saharan Africa.[3] With 14,000 maternal deaths annually and a maternal mortality rate of 412/100,000 live births, Ethiopia is one of six countries^a^ contributing to more than half of global maternal deaths each year.[3-5] Sub-Saharan Africa is the region with the highest under-five mortality, and Ethiopia is one of five countries^b^ that contributed to more than half of the global under-five deaths in 2018.[6] Neonatal mortality has remained stagnant (from 29 to 30/1,000 live births) since 2016.[4,7]

Ethiopia has recorded a significant reduction (71%) in maternal mortality but failed to achieve the Millennium Development Goal (MDG) 5 target of a 75% reduction. The country achieved the MDG 4 of reduction in under-five mortality.[8] However, Ethiopia continues to experience significant maternal and child mortality from preventable causes, such as pregnancy complications (e.g. hypertensive disorders, postpartum hemorrhage, obstetric fistula), complications of preterm birth, and communicable diseases (e.g. lower respiratory infections, gastrointestinal infections). Most maternal deaths (52.5%) occur in the postpartum period and most deaths under five (40.2–54.5% depending on the source) occur in the neonatal period, the first 28 days of life.[7,9,10]

Around the world, populations that have the highest burden of disease also have the least data, research, and programs. Within Ethiopia, more information is needed to understand the current knowledge of maternal, newborn, and child health (MNCH) and identify remaining gaps to prioritize and guide future research. In this scoping review, we aim to synthesize the evidence on MNCH research in Ethiopia over the past 72 years.

## METHODS

The HaSET (“happiness” in Amharic) Maternal and Child Health Research Program conducted a scoping review of the MNCH literature in Ethiopia using a population, concepts, and context (PCC) framework.[11-13] The review was conducted in the following phases: 1) identification of the research question; 2) identification of relevant studies in electronic databases; 3) study selection by screening with inclusion and exclusion criteria; 4) data abstraction; and 5) analysis, interpretation, and synthesis of results.

### Patient and Public Involvement

As this research is a scoping review of the published literature, no patients or the public were involved in the completion of this work.

### Protocol

The protocol for our scoping review was prepared using the Preferred Reporting Items for Systematic reviews and Meta-Analyses extension for Scoping Reviews (PRISMA-ScR) checklist as a guide. The protocol was published in BMJ Open (http://dx.doi.org/10.1136/bmjopen-2019-034307).

### Eligibility criteria

Inclusion criteria was outlined using the PCC framework described by JBI, and is shown in Table 1.[13]

**Table 1:**
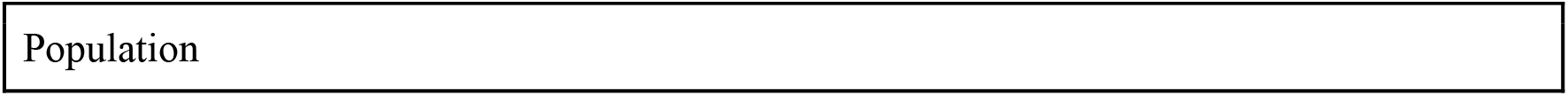
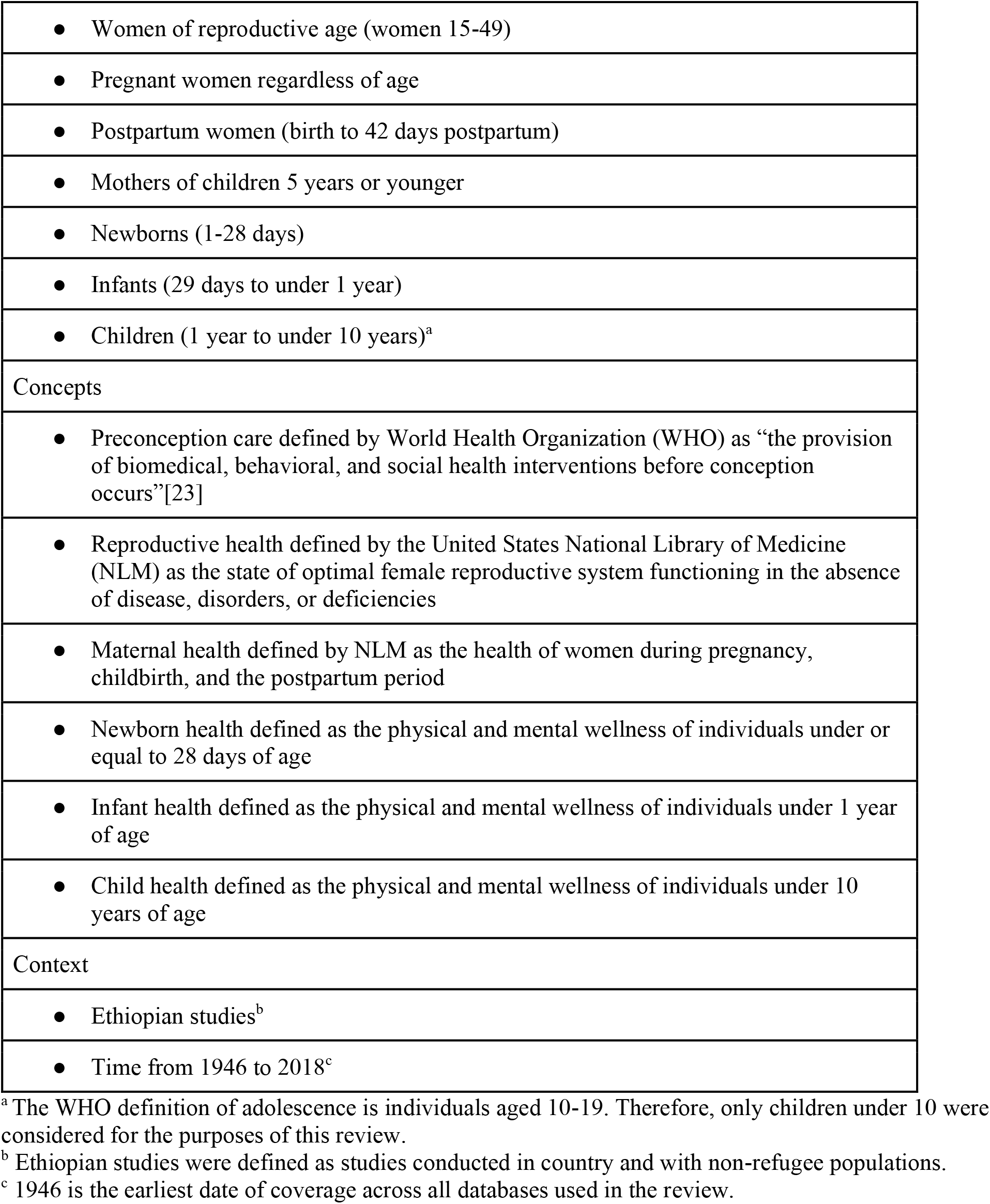
Inclusion criteria using PCC framework

|  |
| --- |
| • Women of reproductive age (women 15-49) |
| • Pregnant women regardless of age |
| • Postpartum women (birth to 42 days postpartum) |
| • Mothers of children 5 years or younger |
| • Newborns (1-28 days) |
| • Infants (29 days to under 1 year) |
| • Children (1 year to under 10 years)<sup>a</sup> |
| Concepts |
| • Preconception care defined by World Health Organization (WHO) as “the provision of biomedical, behavioral, and social health interventions before conception occurs”[23] |
| • Reproductive health defined by the United States National Library of Medicine (NLM) as the state of optimal female reproductive system functioning in the absence of disease, disorders, or deficiencies |
| • Maternal health defined by NLM as the health of women during pregnancy, childbirth, and the postpartum period |
| • Newborn health defined as the physical and mental wellness of individuals under or equal to 28 days of age |
| • Infant health defined as the physical and mental wellness of individuals under 1 year of age |
| • Child health defined as the physical and mental wellness of individuals under 10 years of age |
| Context |
| • Ethiopian studies<sup>b</sup> |
| • Time from 1946 to 2018<sup>c</sup> |

<sup>a</sup> The WHO definition of adolescence is individuals aged 10-19. Therefore, only children under 10 were considered for the purposes of this review.
<sup>b</sup> Ethiopian studies were defined as studies conducted in country and with non-refugee populations.
<sup>c</sup> 1946 is the earliest date of coverage across all databases used in the review.

All evidence-based studies or studies that applied “principles of scientific reasoning, including systematic uses of data and information systems, and appropriate use of behavioral science theory and program planning models” in published literature were included in our review.[14] Examples of such studies included RCTs, observational studies, physiologic studies, case studies, laboratory studies, systematic reviews, and meta-analyses.[15-18]

We excluded studies that were non-English or non-Amharic (the national language of Ethiopia), non-human populations, personal opinion pieces, literature reviews, and not peer reviewed. Studies among sex workers, non-Ethiopian refugee populations, and children over the age of 10 were excluded as outside the scope of our review. We counted the number of studies excluded based on these populations to understand the potential biases in our own review.

### Search strategy

To identify relevant studies, we searched PubMed/MEDLINE, EMBASE, and the WHO African Index Medicus with specific PCC terms (Appendix 1. Search strategy). We conducted the search on 15 January 2019 for entries published from 1946 through 2018. Review of gray literature was not within the scope of this review. The search results were downloaded and imported into a comprehensive library using EndNote version X9.

### Study selection

#### Study selection tools

We developed study selection tools for title and abstract screening and full-text review. Two independent reviewers piloted a random sample of references. The kappa statistic was used to calculate the level of agreement between the two reviewers, and disagreements were discussed before implementing changes to the screening forms. We continued revising the forms until agreement between reviewers was high (kappa statistic > 0.8).

After finalizing screening tools, two research assistants independently screened references for inclusion using the eligibility criteria described in the PCC framework above. This selection process was conducted in two phases: 1) title and abstract screening and 2) full-text review. Any disagreements were resolved through discussion and consensus between the two reviewers. If consensus was unable to be reached, a third reviewer from the research team served as a tie-breaker.

### Title and abstract screening

Each study underwent title and abstract screening. During this phase of selection, titles and abstracts were reviewed to classify each study as ‘yes’ (include), ‘no’ (exclude), or ‘unclear’ (criteria should be further evaluated). All studies marked ‘yes’ or ‘unclear’ were moved to full text review for additional screening. We tallied the reasons for excluding each study marked ‘no.’

### Full-text review

After the completion of title and abstract screening, we conducted a full-text review of remaining studies. During this phase of selection, the full texts of studies initially marked ‘yes’ or ‘unclear’ during the title and abstract screening phase were reviewed for inclusion.

### Data abstraction process

Once screening was complete, a data abstraction tool was developed in Qualtrics XM, a customizable online survey tool, to chart results of the review. The tool was piloted and refined in an iterative process similar to the screening tools, as described above.

Two independent reviewers collected data using the Qualtrics extraction tool. All discrepancies were reconciled through discussion and consensus between the pairs. In the event that consensus could not be reached, a third reviewer resolved the disagreement. For quality control, the team coordinator reviewed a random sample of articles (5%) during the data abstraction process and provided guidance to the team to improve standardization among reviewer pairs.

### Data items

We abstracted data on study characteristics (e.g. publication year, funding, country of corresponding author, setting [academic, community-based, laboratory, etc.], region [urban/rural, specific region], target population, sample size, type of data, study question, study type/design) and interventions, exposures, and outcomes studied in our included studies.

### Analysis and synthesis of results

We collated and summarized included studies, tabulating year of publication; funding; setting and/or region; population; study question and design; and types of interventions, exposures, and outcomes. Analyses were repeated by specific MNCH populations. An assessment of the quality of included studies was outside the scope of this review.

All study endpoints were evaluated via descriptive analysis and reported in descriptive tables.

## RESULTS

### Selection of studies

Our search strategy identified 8,149 articles, of which 7,820 were unique articles and reviewed. During title and abstract screening, 4,231 articles were excluded. We excluded another 1,419 in the full-text screening. Overall, 3,229 did not include a MNCH population or objective, 684 included children ≥ 10 years of age, 669 were not based in Ethiopia, 378 were conference abstracts, 321 were not evidence-based, 223 were unable to have the full-text version retrieved, 53 were opinion pieces, 51 were not human-based studies, 26 included sex workers, 13 included non-Ethiopian refugee populations, and three were not published in English. The remaining 2,170 studies were considered eligible and included in our final review. Figure 1 illustrates the PRIMSA flow diagram of studies through the different stages of the scoping review, mapping out the number of records identified, included, excluded, and reasons for exclusion.

**Figure 1:**
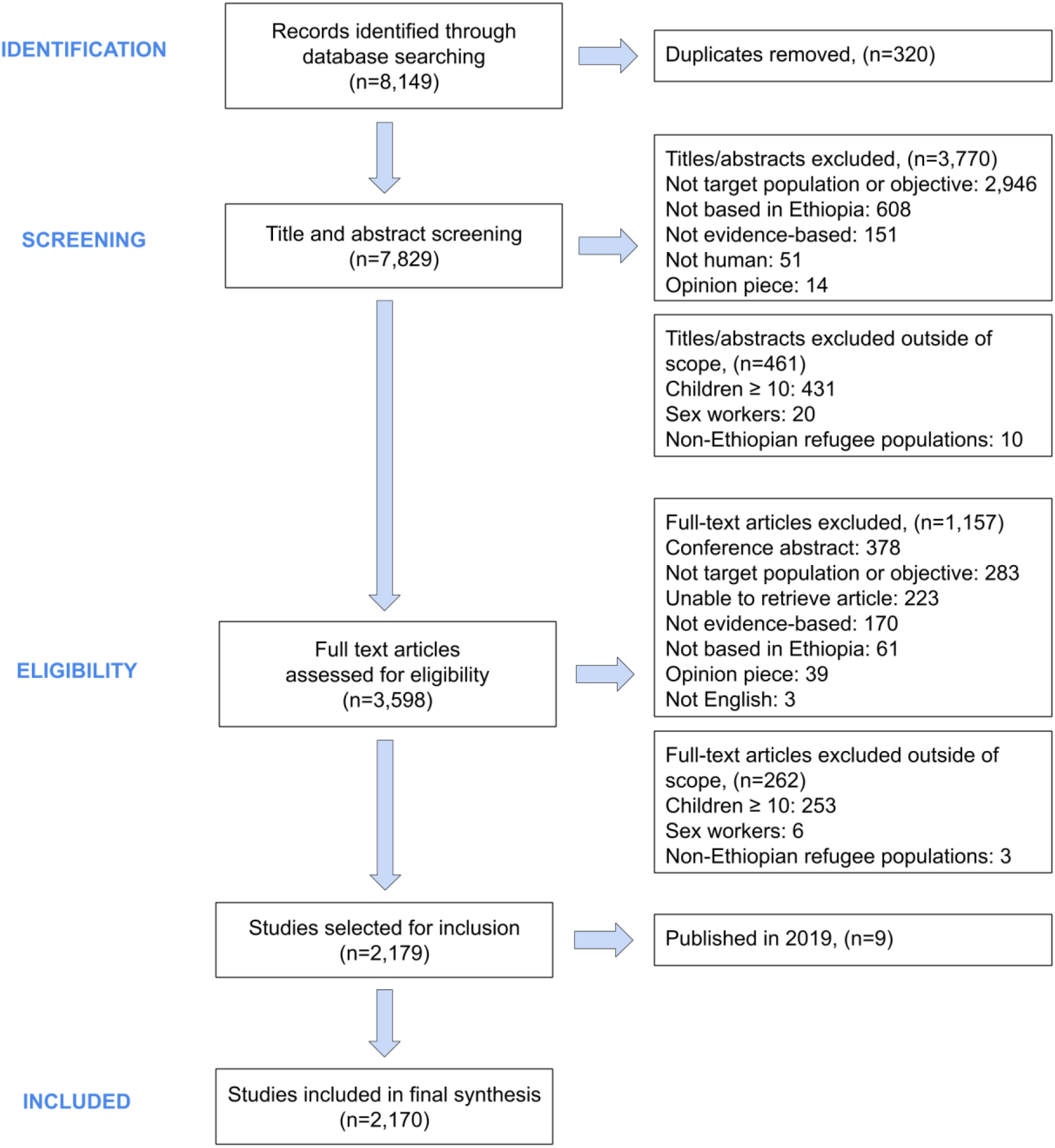
Flow diagram of inclusion process

From 1946 to 2018, the number of published studies (n=2,170) on MNCH in Ethiopia increased (Figure 2) with a significant percentage of that increase in recent years. Indeed, the majority of studies were published after 2000 (n=1,823, 84.0%), with 1,518 (70.0%) of total studies published within the past decade.

**Figure 2:**
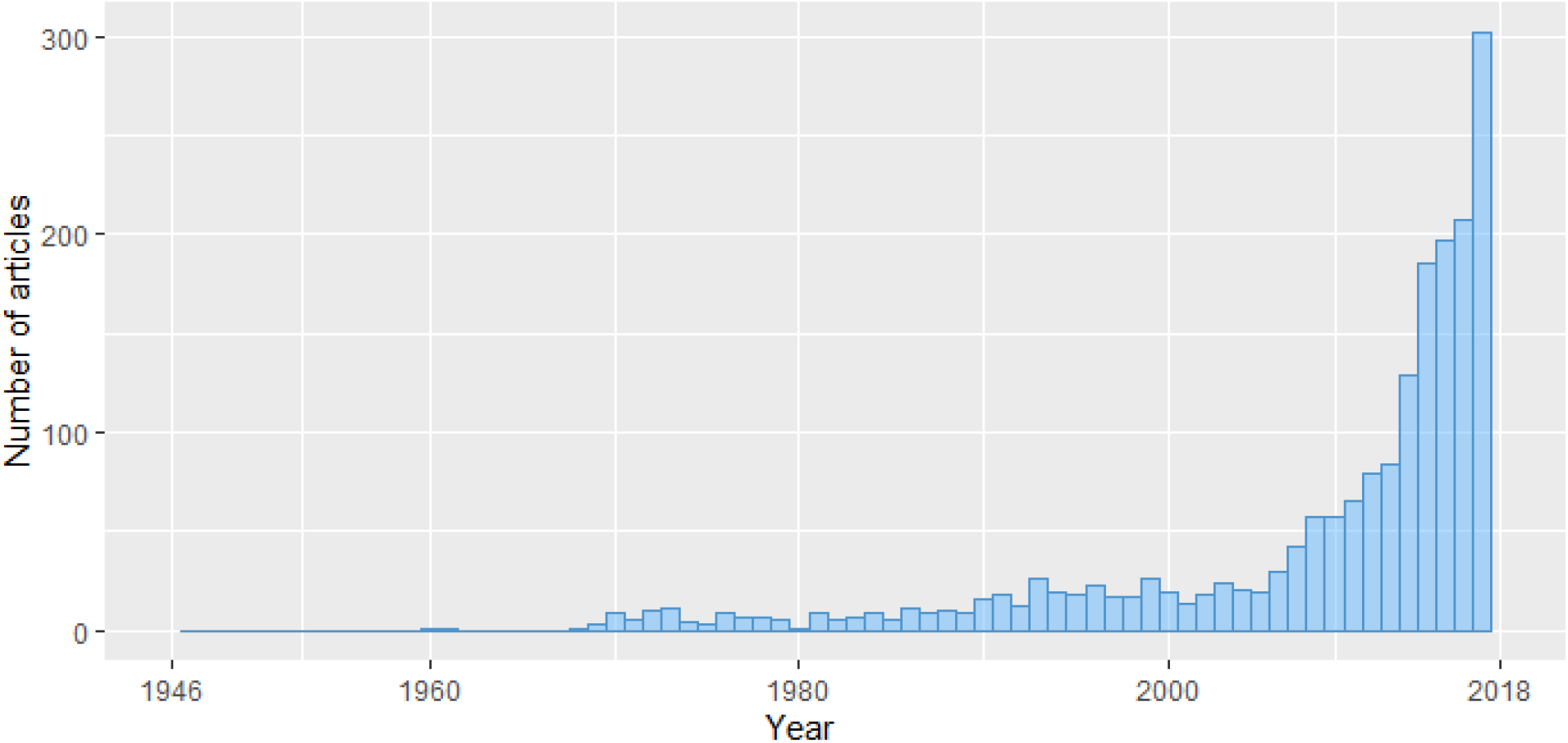
Distribution of publications by year

### Study settings and populations

The majority of studies (n=1,984, 91.4%) were single-country studies focused on Ethiopia. Of the 1,869 reporting a geographic region of study, 1,672 (89.5%) were conducted within Addis Ababa (n=419, 22.4%) and the four agrarian regions (Amhara [n=367, 19.6%], Oromia [n=341, 18.2%], Southern Nations, Nationalities, and Peoples’ Region [n= 338, 18.1%], and Tigray [n=207, 11.1%]), as compared to the pastoralist and semi-pastoralist regions (n=197, 10.5%). Households were the most common study setting, with 46.2% of studies (n=1,064) conducted as household surveys. Other study settings included health facilities (n=741, 32.2%) such as health posts, health centers, or district hospitals not affiliated with an academic institution; and academic-related health facilities (n=304, 13.2%) such as university hospitals.

Of the 1,218 studies (56.1%) that defined urban, rural, and mixed (both urban and rural) settings, 751 (61.7%) included populations in a mixed setting, 339 (27.8%) included populations in a rural setting, and 128 (10.5%) included populations in an urban setting. Of the studies that reported a mixed residence, the participants predominantly lived in a rural setting (63.1%) compared to 36.9% living in an urban area.

The most common MNCH populations studied were children aged one to less than 10 years (n=765, 30.5%), women of reproductive age (n=552, 22.0%), and pregnant women (n=548, 21.9%). The least studied population was postpartum women up to six weeks post birth (n=72, 2.9%) (Figure 3).

**Figure 3:**
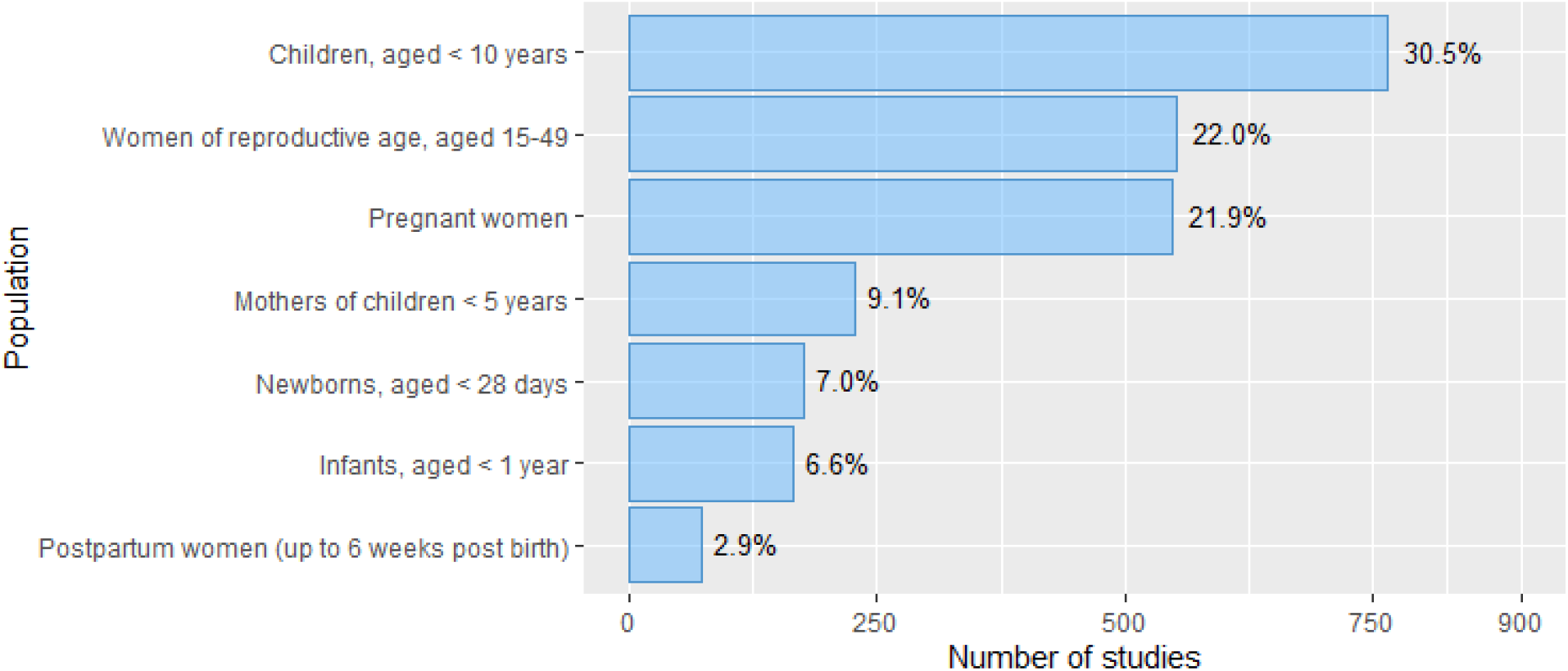
Number of studies published by target population Note: selection of target populations was not mutually exclusive

### Study characteristics

On study design, the majority of the studies (n=1,298, 53.6%), were cross-sectional. Qualitative studies (n=138, 5.7%), prospective cohort studies (n=134, 5.5%), and case-control studies (n=125, 5.2%) were less common. Systematic reviews (n=45, 1.9%) and meta-analyses (n=28, 1.2%) were rare. RCTs (n=56) (clustered, individual, and block) accounted for less than 3% of our total included studies.

The most frequently reported types of research questions were association/etiology (n=1,284, 37.1%), prevalence (n=875, 25.3%), and descriptive (n=613, 17.7%), consistent with the high proportion of cross-sectional studies. The least reported types of research questions were cost-effectiveness (n=25, 0.7%), screening (n=23, 0.7%), and prognosis, or the course of an illness over time (n=22, 0.6%).

Most included studies (n=1,889, 87.1%) analyzed quantitative data. Of the remaining studies, 131 studies (6.0%) used qualitative data and 144 (6.6%) used both quantitative and qualitative data. The median length of included studies was five months (IQR: 2-17). The median sample size of included studies was 470 individuals (IQR: 206-1,122.5). See Appendix 2. Types of study design.

In more than half (n=1,399, 64.5%) of the included studies, the corresponding author was from Ethiopia. The next most frequently reported country of origin for corresponding authors included the United States of America (n=284, 13.1%), the United Kingdom (n=97, 4.5%), and Sweden (n=50, 2.3%).

On funding, 1,257 studies (57.9%) reported receipt of funding. Of these funded studies, 457 (36.4%) included children aged one to less than 10 years, 320 (25.5%) included pregnant women, and 302 (24%) included women of reproductive age. The largest source of funding was from domestic (Ethiopian) academic institutions (n=479, 29.4%). Federal foreign agencies (e.g. USAID) and non-UN international agencies (e.g. Bill & Melinda Gates Foundation) were also frequently reported, funding 350 (21.5%) and 328 (20.2%) studies, respectively.

### Research topics studied

To describe the scope of research topics among studies with quantitative data (n=1,929, 88.9%), we present the data by studies that included interventions, exposures, and/or outcomes data. See Appendix 3. Summary of research topics for a table of research topics.

#### Interventions

Of the total number of included studies, 9.2% (n=200) evaluated interventions. Among children, 34 studies (22.2%) tested non-HIV, non-malaria, non-diarrheal infectious disease prevention and/or management interventions, 18 (11.8%) tested diarrheal disease prevention/management, and 17 (11.1%) tested malaria prevention/management. Two studies (1.3%) evaluated management for those exposed to or infected with HIV. Twenty-one studies (13.7%) included malnutrition management interventions.

Among infants, the most commonly studied interventions were related to nutrient supplementation (n=8, 20.5%) of which four (50%) included zinc supplementation, two (25%) included general micronutrient supplementation, one (0.13%) included omega-3 fatty acid supplementation, and one (0.13%) included vitamin A supplementation. Additional interventions included diarrheal disease prevention/management (n=5, 12.8%), malnutrition management (n=4, 10.3%), and management for those exposed to or infected with HIV (n=4, 10.3%).

Among newborns, the most frequently reported interventions were antibiotic prophylaxis or treatment (n=5, 16.7%). Of the five studies, two studies (40%) evaluated injectable gentamicin and oral amoxicillin for the treatment of bacterial infection; three (60%) mentioned use for prevention or management of newborn infection, newborn sepsis, and/or pneumonia. Additionally, two studies (11.1%) evaluated management for newborns exposed to or infected with HIV.

While interventions on newborns, infants, and children primarily focused on infectious diseases or nutrition management, interventions on the maternal population (pregnant women, postpartum women, or mothers of children less than five years of age) and women of reproductive age were more frequently centered around healthcare utilization and strengthening, health promotion, and reproductive health. In the maternal population, most interventions studied (n=20, 23.8%) were related to health system strengthening. Examples of these interventions include training programs for healthcare providers or community members (n=9, 45%), improving referral networks and capacity (n=6, 30%), and implementation of the Health Extension Program (n=3, 15%). Other notable interventions for the maternal population involved health or healthcare promotion (n=12, 14.3%) and labor and delivery management (n=10, 11.9%). Among studies with women of reproductive age, family planning interventions (n=17, 47.2%) were most common.

#### Exposures

Most studies, 1,466 (67.6%), investigated one or more exposures. The most common exposures were demographics and social determinants of health (n=1,318, 43.4%), nutrition (n=463, 15.2%), and infectious disease (n=394, 13.0%). Accidents and injury (n=4, 0.1%) were the least common exposure. Within demographics and social determinants of health, the most frequently reported responses were related to age (n=1,028, 13.5%); education (n=891, 11.7%); and health, healthcare, and health seeking behaviors (n=690, 9.1%).

#### Outcomes

Of the 875 outcomes reported for pregnant women, postpartum women, or mothers of children less than five years of age, the main outcomes studied included health service utilization (n=266, 30.4%), infectious pregnancy complications (n=151, 17.3%), and nutrition (n=133, 15.2%). The least reported outcomes for the maternal population was violence (n=6, 0.7%).

Outcomes reported for children (n=844) were most frequently related to infectious disease (n=305, 36.1%), nutrition (n=256, 30.3%), and mortality (n=105, 12.4%). Psychosocial and violence and injury outcomes were disproportionately lower, at 1.3% (n=11) and 0.6% (n=5), respectively.

Infant outcomes (n=189) were similar to those reported for children, with outcomes distributed among infectious disease (n=62, 32.8%), nutrition (n=58, 30.7%), and mortality (n=39, 20.6%). However, no psychosocial or violence and injury outcomes were reported for infants. The most frequently reported outcome for newborns (n=415) was mortality (n=108, 26%).

Women of reproductive age outcomes (n=404) primarily focused on family planning (n=161, 39.9%). Other frequently reported outcomes were distributed across infectious disease (n=49, 12.1%), psychosocial (n=29, 7.2%), violence and injury (n=27, 6.7%), and nutrition (n=26, 6.4%).

## DISCUSSION

To our knowledge, this is the first study to describe the broad and diverse landscape of the maternal and child health literature in Ethiopia. Our findings show a significant upward trend in MNCH research interest since 2000. The increase may be related to commitment from Ethiopian institutions and the Ministry of Health to improve MNCH health outcomes. The rapid expansion of over 40 universities and health science colleges in the past two decades has contributed to a growing body of motivated young researchers in Ethiopia.

Despite the increase in research across the past two decades, we found a gap in research activities among certain populations (newborns and postpartum women). More than three quarters of the studies focused on children less than 10 years of age and women of reproductive age (pregnant or non-pregnant). Study types were limited to simple cross-sectional designs and focused on descriptive analyses. The most frequently reported study questions included associations/etiologies, prevalence calculations, and descriptive characteristics, with very few studies focused on evaluating interventions, prevention measures, and screening of MNCH populations. Almost half of all exposures reported were demographics and social determinants of health (e.g. age, economic stability, education). Infectious outcomes were most frequently reported for children, while those for the maternal population and women of reproductive age primarily focused on utilization of health services such as antenatal and postnatal care and family planning. Since Ethiopia has the sixth highest number of maternal deaths globally and fourth highest number of newborn deaths, additional studies beyond utilization of health services and descriptions of demographics and social determinants are needed to improve risk prediction and identification of high risk conditions, investigate mechanisms of preterm and stillbirths to develop and test interventions, and empower individuals and families to make decisions and advocate for their health care.[5,19]

Most studies were conducted in Addis Ababa and/or the four agrarian regions, whereas pastoralist and semi-pastoralist regions experience higher morbidity and mortality rates.[20] Although Addis Ababa only contributes to 4% of the total population, more than 20% of the studies were conducted in Addis Ababa. Pastoralist regions account for approximately 14% of the population and 61% of total geographic area, but were represented in less than 10% of the studies.[21] Pastoralist populations are frequently mobile, lack access to infrastructure in remote areas, lack access to basic services (e.g. education, healthcare), hold strong traditional and cultural beliefs, and are consequently difficult to engage. These populations remain underrepresented in research and household surveys.[22] Along with challenges brought on by nomadic life, there are logistical challenges to conducting research in remote areas with limited infrastructure.

Through this review, we identified significant gaps in the body of MNCH research in Ethiopia. Postpartum women up to six weeks post birth and newborns were neglected populations in the MNCH research studies. Psychosocial exposures represented a small fraction of all exposures reported in included studies, and no interventions on mental health were captured among any population. Across all MNCH populations, violence and injury were neglected areas of research. Although the postpartum period is a short timespan across a lifetime, it represents a period when both mothers and their children die at a higher rate. Specific data from studies that examine the effect of psychosocial exposure, mental health, and violence and injuries may drive interventions that can improve national statistics on MNCH mortality and advance the health of children while protecting mothers at the same time.

### Limitations

The results of this review focused primarily on Ethiopia and may not be generalizable to other countries. However, this study may inform the need to conduct similar landscape reviews in all sub-Saharan countries and across religious or ethnic groups. The targeted geographic region will support prioritization of research questions, study designs, and study population for future research. Due to the extensive number of studies included across diverse subject matters within MNCH, the quality of each study was not assessed. Additional work on quality assessment of studies may be conducted in the future, although assessment of methodologic quality will be limited because most studies were cross-sectional.

Additionally, the scoping review was limited to published data in indexed journals, leaving our review subject to the effects of publication bias. Many studies conducted on MNCH in Ethiopia are published in local journals or available only as hard copies that are not indexed in international databases and are consequently not reflected in our review.

## CONCLUSION

To our knowledge, this is the first scoping review to describe the complete landscape of MNCH research in Ethiopia. Understanding the depth of existing MNCH knowledge in Ethiopia is critical for the prioritization and development of future research questions. The results of this review have allowed us to identify prominent gaps in the research and have underscored the need for additional studies on the postpartum period; psychosocial health; and causes and effects of violence, accidents, and injury on all MNCH populations. Globally in low and middle-income country settings, these are neglected areas of research but may contribute to morbidity and mortality. In consultation with key in-country stakeholders, these results will be disseminated and used to inform future priority research questions within the field. Moving forward, a shift of focus towards gold standard methodology, particularly RCTs, is needed to build a more rigorous evidence base for the translation of research to policy and improvement of overall health for mothers and children in Ethiopia.

## Supporting information

Appendix 1. Search strategy

## Data Availability

All data relevant to the study are included in the article or can be requested from the authors.

## CONTRIBUTORS

GJC conceived the idea and study design and contributed to the data analysis, interpretation of findings, and writing. JD contributed to the writing, data abstraction, data analysis and interpretation, and development of figures and appendices. MG, MK, RO abstracted data and contributed to the study design, data analysis, and interpretation. BH, SU, LT, DB contributed to the writing and data interpretation. All authors approved the final version of the paper for submission.

## ACKNOWLEDGEMENTS

We appreciate Alexandra Rashedi, Anthony Sun, Cristina Alonso, Francesca Deleso-Frechette, Hemen Muleta, Ismaeel Yunusa, Jordan Arvayo, Jose Gonzalez, Melie Anyachebelu, Michelle Olakkengil, Mirzya Haider, Moria Mahanaimy, Muhummed Nadeem Kasmani, Neena Kapoor, Purvaja Kavattur, Quinn McVeigh, Ramya Pinnamaneni, and Thein Min Swe for contributing to the data abstraction process. We thank Ebba Abate, Getachew Tollera, and Theodros Getachew for their support and commitment to our work at the Ethiopian Public Health Institute.

## COMPETING INTERESTS

None declared.

## FUNDING

This work was supported by the Bill & Melinda Gates Foundation grant number OPP1201842.

## Footnotes

a The six countries contributing to more than half of all maternal deaths in 2008 were India, Nigeria, Pakistan, Afghanistan, Ethiopia, and the Democratic Republic of the Congo.

b The five countries contributing to half of global under-five deaths in 2018 were India, Nigeria, Pakistan, Democratic Republic of the Congo, and Ethiopia.

## Notes

### Competing Interest Statement

The authors have declared no competing interest.

### Author Declarations

Research ethics approval was not applicable because the study did not involve human participants.

