## Appendix 1. Search strategy for "Gaps in maternal, newborn, and child health research: a scoping review of 72 years in Ethiopia"

Detailed search strategy for Ethiopian MNCH research scoping review
Last updated January 15, 2019

### PUBMED

| | |
|---|---|
| *Population* | (("Mothers"[Mesh] OR mother*[tiab] OR matern*[tiab] OR women*[tiab] OR "reproductive age"[tiab] OR "Pregnant Women"[Mesh] OR "Pregnancy"[Mesh] OR pregnan*[tiab] OR gravid*[tiab] OR prenatal[tiab] OR antenatal[tiab] OR pre natal[tiab] OR ante natal[tiab] OR perinatal[tiab] OR postnatal[tiab] OR post natal[tiab] OR postpartum[tiab] OR "Peripartum period"[Mesh] OR "Postpartum period"[Mesh]) |
| | OR |
| | ("Infant"[Mesh] OR infant*[tiab] OR newborn[tiab] OR neonat*[tiab] OR fetal[tiab] OR fetus[tiab] OR feotal[tiab] OR feotus[tiab] OR "Child"[Mesh] OR child*[tiab] OR boy[tiab] OR boys[tiab] OR girl[tiab] OR girls[tiab] OR toddler*[tiab] OR "Adolescent"[Mesh] OR teen*[tiab] OR adolescen*[tiab] OR youth[tiab]) |
| OR | |
| *Concepts* | ("Reproductive Health"[Mesh] OR "Reproductive Behavior"[Mesh] OR "Reproductive History"[Mesh] OR "Contraception"[Mesh] OR "Family Planning Services"[Mesh] OR "family planning"[tiab] OR "birth control"[tiab] OR contraception[tiab] OR "Birth Intervals"[Mesh] OR "Reproductive Health Services"[Mesh] OR "Maternal Health Services"[Mesh] OR "Child Health Services"[Mesh] OR "Maternal-Child Health Centers"[Mesh] OR "Maternal-Child Nursing"[Mesh] OR "Obstetric Nursing"[Mesh] OR "Pediatric Nursing"[Mesh] OR "Doulas"[Mesh] OR "Midwifery"[Mesh] OR "Nurse Midwives"[Mesh] OR doula*[tiab] OR midwife*[tiab] OR "Obstetric Surgical Procedures"[Mesh] OR "Prenatal Care"[Mesh] OR "Prenatal Education"[Mesh] OR "Hospitals, Maternity"[Mesh] OR "Diagnostic Techniques, Obstetrical and Gynecological"[Mesh] OR "Maternal Exposure"[Mesh] OR "Maternal Age"[Mesh] OR "Infectious Disease Transmission, Vertical"[Mesh] OR "Postnatal Care"[Mesh] OR "Kangaroo-Mother Care Method"[Mesh] OR "Intensive Care Units, Pediatric"[Mesh] OR "Intensive Care, Neonatal"[Mesh] OR "Maternal Nutritional Physiological Phenomena"[Mesh] OR "Prenatal Nutritional Physiological Phenomena"[Mesh] OR "Child Nutritional Physiological Phenomena"[Mesh] OR "Infant Nutritional Physiological Phenomena"[Mesh])<br>OR<br>("Maternal Health"[Mesh] OR "maternal health"[tiab] OR "Maternal Welfare"[Mesh] OR "mother-child"[tiab] OR "Mother-Child Relations"[Mesh] OR "Maternal Behavior"[Mesh] OR "Maternal Mortality"[Mesh] OR "Parturition"[Mesh] OR birth[tiab] OR "Pregnancy Complications"[Mesh] OR "Genital Diseases, Female"[Mesh] OR "Pelvic Floor Disorders"[Mesh] OR "Gestational Weight Gain"[Mesh] OR "Birth Weight"[Mesh] OR "childbirth"[tiab] OR "childbirth complications"[tiab] OR "Depression, Postpartum"[Mesh] OR "Postpartum Hemorrhage"[Mesh] OR "Infertility, Female"[Mesh] OR "Fertility"[Mesh] OR abortion*[tiab] OR miscarriage*[tiab] OR stillbirth*[tiab] OR "Congenital, Hereditary, and Neonatal Diseases and Abnormalities"[Mesh] OR "Infant Health"[Mesh] OR "Infant Mortality"[Mesh] OR "Child Mortality"[Mesh] OR "Neurodevelopmental Disorders"[Mesh] OR "Child Behavior Disorders"[Mesh] OR "Child Behavior"[Mesh] OR "Child Development"[Mesh] OR "Child Welfare"[Mesh] OR "Child Health"[Mesh] OR "Adolescent Health"[Mesh] OR "Adolescent Development"[Mesh])) |
| AND | |
| *Context* | ("Ethiopia"[Mesh] OR ethiopia[tiab]) |
| Filters | English, Human |

Number of results: 5896
Last searched: January 15, 2019

### EMBASE

| | |
|---|---|
| *Population* | (('adolescent mother'/de OR 'expectant mother'/de OR 'surrogate mother'/exp OR 'mother*':ab,ti OR 'matern*':ab,ti OR 'women*':ab,ti OR 'reproductive age':ab,ti OR 'named groups by pregnancy'/exp OR 'pregnancy'/exp OR 'pregnan*':ab,ti OR 'gravid*':ab,ti OR 'prenatal':ab,ti OR 'antenatal':ab,ti OR 'pre natal':ab,ti OR 'ante natal':ab,ti OR 'perinatal':ab,ti OR 'postnatal':ab,ti OR 'post natal':ab,ti OR 'postpartum':ab,ti OR 'perinatal period'/de OR 'puerperium'/de) |
| | OR |
| | ('infant'/exp OR 'infant*':ab,ti OR 'newborn':ab,ti OR 'neonat*':ab,ti OR 'fetal':ab,ti OR 'fetus':ab,ti OR 'feotal':ab,ti OR 'feotus':ab,ti OR 'child'/exp OR 'child*':ab,ti OR 'boy':ab,ti OR 'boys':ab,ti OR 'girl':ab,ti OR 'girls':ab,ti OR 'toddler*':ab,ti OR 'adolescent'/exp OR 'teen':ab,ti OR 'adolescen*':ab,ti OR 'youth':ab,ti) |
| OR | |
| *Concepts* | ('reproductive health'/de OR 'reproductive behavior'/de OR 'reproductive history'/de OR 'contraception'/exp OR 'family planning'/de OR 'family planning':ab,ti OR 'birth control':ab,ti OR 'contraception':ab,ti OR 'maternal health service'/de OR 'maternal child health care'/de OR 'child health care'/exp OR 'newborn nursing'/exp OR 'nurse midwifery'/de OR 'obstetrical nursing'/de OR 'pediatric nursing'/exp OR 'perinatal nursing'/de OR 'midwife'/de OR 'doula'/de OR 'traditional birth attendant'/de OR 'doula*':ab,ti OR 'midwife*':ab,ti OR 'obstetric operation'/exp OR 'prenatal care'/exp OR 'childbirth education'/exp OR 'prepregnancy care'/de OR 'perinatal care'/exp OR 'intrapartum care'/de OR 'maternal exposure'/de OR 'maternal age'/de OR 'vertical transmission'/de OR 'postnatal care'/exp OR |

| | |
|---|---|
| | 'kangaroo care'/de OR 'pediatric intensive care unit'/de OR 'neonatal intensive care unit'/de OR 'newborn intensive care'/de OR 'maternal nutrition'/de OR 'child nutrition'/exp OR 'adolescent nutrition'/de)<br>OR<br>('maternal welfare'/de OR 'maternal health':ab,ti OR 'mother-child':ab,ti OR 'mother child relation'/de OR 'maternal behavior'/de OR 'maternal mortality'/de OR 'birth'/de OR 'birth':ab,ti OR 'pregnancy complication'/exp OR 'gynecologic disease'/exp OR 'gestational weight gain'/de OR 'birth weight'/exp OR 'childbirth':ab,ti OR 'childbirth complications':ab,ti OR 'postnatal depression'/de OR 'postpartum hemorrhage'/de OR 'female infertility'/exp OR 'female fertility'/de OR 'abortion*':ab,ti OR 'miscarriage*':ab,ti OR 'stillbirth*':ab,ti OR 'newborn disease'/exp OR 'infant disease'/exp OR 'child health'/de OR 'infant mortality'/de OR 'developmental disorder'/exp OR 'child behavior'/exp OR 'child development'/de OR 'motor development'/de OR 'child welfare'/exp OR 'adolescent health'/de OR 'adolescent development'/de OR (('adolescent mother'/de OR 'expectant mother'/de OR 'surrogate mother'/exp OR 'mother*':ab,ti OR 'matern*':ab,ti OR 'women*':ab,ti OR 'reproductive age':ab,ti OR 'named groups by pregnancy'/exp OR 'pregnancy'/exp OR 'pregnan*':ab,ti OR 'gravid*':ab,ti OR 'postnatal':ab,ti OR 'post natal':ab,ti OR 'postpartum':ab,ti OR 'perinatal period'/de OR 'puerperium'/de OR 'infant'/exp OR 'infant*':ab,ti OR 'newborn':ab,ti OR 'neonat*':ab,ti OR 'fetal':ab,ti OR 'fetus':ab,ti OR 'feotal':ab,ti OR 'feotus':ab,ti OR 'child'/exp OR 'child*':ab,ti OR 'boy':ab,ti OR 'boys':ab,ti OR 'girl':ab,ti OR 'girls':ab,ti OR 'toddler*':ab,ti OR 'adolescent'/exp OR 'teen*':ab,ti OR 'adolescen*':ab,ti OR 'youth':ab,ti) AND ('mental disease'/exp OR 'behavior disorder'/exp)))) |
| AND | |
| *Context* | ('Ethiopia'/de OR 'ethiopia':ab,ti) |
| Filters | [humans]/lim AND [english]/lim NOT [medline]/lim |

Number of results: 1873
Last searched: January 15, 2019

**WHO AFRICAN INDEX MEDICUS *(via WHO Global Index Medicus)***

**Maternal Health**
(mother* OR matern* OR women* OR "reproductive age" OR pregnan* OR gravid* OR prenatal OR antenatal OR “pre natal” OR “ante natal” OR perinatal OR postnatal OR “post natal” OR postpartum OR peripartum OR “maternal health” OR “reproductive health”) AND (Ethiopia)

Limit: English
Number of results: 137
Last searched: January 15, 2019

**Newborn and Infant Health**
(infant* OR newborn OR neonat* OR fetal OR fetus OR feotal OR feotus OR “infant health” OR “newborn health” OR birth) AND (Ethiopia)

Limit: English
Number of results: 150
Last searched: January 15, 2019

**Child and Adolescent Health**
(child* OR boy OR boys OR girl OR girls OR toddler* OR adolescent OR teen* OR adolescen* OR youth) AND (Ethiopia)

Limit: English
Number of results: 93
Last searched: January 15, 2019

Total number of MNCH results from AIM: 380

Total number of results across three databases as of January 15, 2019: 8149
